# A Snapshot of COVID-19 Incidence, Hospitalizations, and Mortality from Indirect Survey Data in China in January 2023

**DOI:** 10.1101/2023.02.22.23286167

**Authors:** Juan M. Ramírez, Sergio Díaz-Aranda, Jose Aguilar, Oluwasegun Ojo, Rosa Elvira Lillo, Antonio Fernández Anta

## Abstract

In this work we estimate the incidence of COVID-19 in China using online indirect surveys (which preserve the privacy of the participants). The indirect surveys deployed collect data on the incidence of COVID-19, asking the participants about the number of cases, deaths, vaccinated, and hospitalized that they know. The incidence of COVID-19 (cases, deaths, etc.) is then estimated using a modified Network Scale-up Method (NSUM). Survey responses (100, 200 and 1,000, respectively) were collected from Australia, the UK, and China in January 2023. The estimates in Australia and the UK are compared with official data, showing that they are in the confidence intervals or rather close. Cronbach’s alpha values also indicate good confidence in the estimates. The estimates obtained in China are, among others, that 91% of the population is vaccinated, almost 80% had been infected in the last month, and almost 3% in the last 24 hours.

## 1 Introduction

Being able to have estimates of the incidence of COVID-19 on variables such as the number of cases, deaths, and hospitalizations, among others, is of great interest to decision-makers. Direct surveys have been used in several countries during COVID-19 to estimate these variables [1, 2]. Unfortunately, these surveys need large numbers of participants to achieve reliable estimates and usually collect sensitive personal data (which may deter respondents due to privacy concerns and require careful data manipulation).

As an alternative to these surveys, there are indirect surveys, in which the questions answered by the participants are not about themselves, but about their contacts. Estimates are then obtained from indirect survey responses using the *Network Scale-up Method* (NSUM) [3, 4]. This approach allows 1) reaching a larger sub-population, 2) reducing data collection cost, and 3) estimating the true value with a computationally efficient method, and 4) provide high privacy for participants. Indirect surveys, have already been used for COVID-19 [5].

We use indirect online surveys to estimate cases, mortality, vaccinated, and hospitalizations due to COVID-19 in China for the period of January 18-26, 2023. We validate our approach using data from Australia and the United Kingdom (UK) collected on January 19, 2023. These metrics are compared with the official values reported by Our World in Data (OWID) and the Office for National Statistics (ONS) from UK. In addition, we use Cronbach’s alpha index [6], which is a reliability value to measure the internal consistency of the questionnaire generated by indirect surveys.

## 2 Methods

### 2.1 Sampling Participants

We conducted online indirect surveys using the PollFish platform. Specifically, we conducted an online survey in China between January 18-26, 2023. This survey collected information about various COVID-19 indicators (vaccination, deaths, and number of cases in the last month, the last 7 days, and the past 24 hours) among the 15 closest contacts of 1,000 participants (see Supplementary Information section for the Chinese and English versions of the survey questions). Additionally, for validation, we conducted online surveys in Australia (100 responses) and the UK (200 responses) on January 19, 2023. Table 3 in Supplementary Information shows the characteristics of the survey respondents (the platform provides information on gender, age group, education, and ethnicity). The respondents of each survey are also stratified by region. For instance, Fig. 1 in Supplementary Information shows a map of China where the intensity corresponds to the number of questionnaires completed in each province.

**Table 1:** COVID-19 incidence metrics in % (and 95% CI) obtained from indirect survey data and official reports for Australia and the UK. (1) People aged 12 years and over that have received at least one/two/three doses on Aug 31, 2022. (2) England data only, 5 weeks.

|  | Australia |  | United Kingdom |  |  |
| --- | --- | --- | --- | --- | --- |
|  | Indirect Survey | OWID | Indirect Survey | OWID | ONS |
| Cases (last month) | 12.43 (10.26 - 14.60) | 1.585 | 8.67 (7.39 - 9.96) | 0.305 | 9.663 |
| Vaccination rate | 76.50 (73.70 - 79.29) | 84.95 | 78.86 (77.00 - 80.72) | 79.71 | 93.6/88.2/70.2<sup>(1)</sup> |
| Mortality (last month) | 0.34 (0.00 - 0.72) | 0.006 | 0.43 (0.13 - 0.73) | 0.004 | 0.005<sup>(2)</sup> |
| Hospitalizations (last month) | 1.02 (0.36 - 1.68) | 1.327 | 0.81 (0.40 - 1.22) | 0.158 | 0.044<sup>(2)</sup> |
| Cases (24 hours) | 2.03 (1.10 - 2.96) | 0.069 | 1.30 (0.78 - 1.82) | 0.023 | 1.458 |
| Cases (7 days) | 2.71 (1.64 - 3.78) | 0.211 | 1.30 (0.78 - 1.82) | 0.073 | 1.116 |
| Cronbach's alpha | 0.83 |  | 0.95 |  |  |

**Figure 1:**
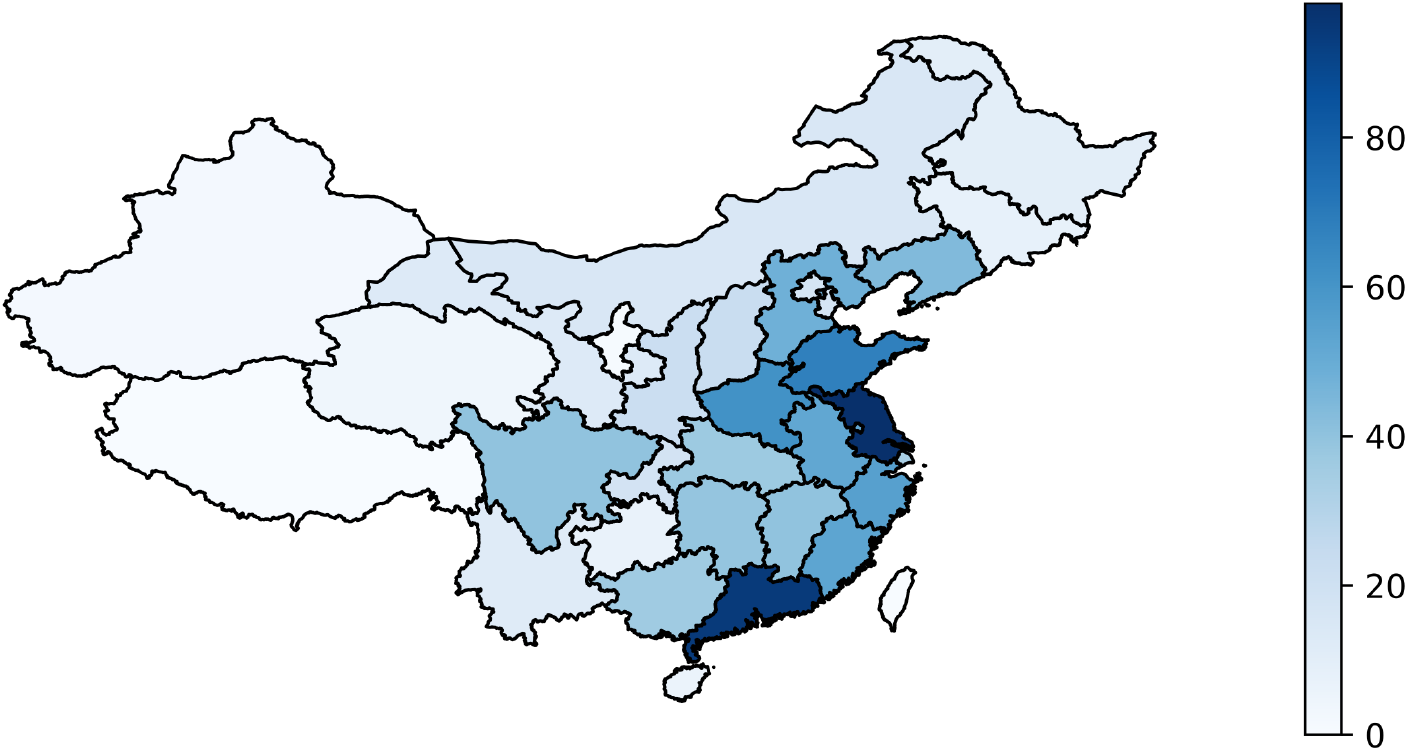
Number of completed questionnaires for the survey deployed in China

### 2.2 Data Analysis

In order to obtain a reliable dataset, we performed two subphases of preprocessing: (1) an inconsistency filter, and (2) a univariate outlier detection.

1. The inconsistency filter removes participants with inconsistent responses: less infected contacts than fatalities, less infected contacts than hospitalized, less infected contacts in the last month than in the last 7 days, and less infected contacts in the last month than in the last 24 hours.
2. Since the collected variables exhibit extremely skewed distributions, the robust outlier detection method reported in [7] is applied. Based on the variable data, this method firstly estimates the quartiles *Q*_1_ and *Q*_3_, as well as the interquartile range (*IQR*). Then, the whiskers *Q*_*α*_ and *Q*_*β*_ are set. Finally, this method preserves the samples in the interval limited by

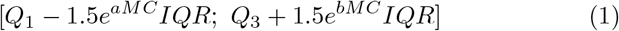

where *MC* is the medcouple statistic that estimates the degree of skewness of the data. Samples outside the interval are marked as outliers and, consequently, are removed. In addition, to estimate the parameters *a* and *b*, we consider the system [7]

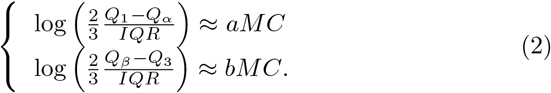

To estimate cumulative incidences, hospitalization rates, and mortality rates, we use a modification of NSUM. Let *c*_*i*_ be the number of contacts of *i*-th respondent with a particular characteristic, e.g., persons who are hospitalized. Further, consider *r*_*i*_ the number of close contacts of the *i*-th respondent (which in this study is fixed at *r*_*i*_ = 15, as shown in the questions in the Supplementary Information). The requirement of close contacts is introduced to minimize the effect of the visibility bias [8] with respect to the classical method [3]. We estimate the aggregated rate, *p*, as ∑_*i*_ *c*_*i*_*/* ∑_*i*_ *r*_*i*_ = ∑_*i*_ *c*_*i*_*/*(15*n*). The estimator’s variance is 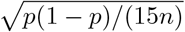, assuming that the *c*_*i*_ are independent binomial random variables with 15 trials and success probability *p*.

To determine the validity of our method, we compared the difference between the official values reported in *Our World in Data* (OWID)^1^ and the values estimated in our approach, for Australia and UK (see Table 1). For both countries, official data were extracted between December 20, 2022, and January 19, 2023. To transform hospital occupancy into the number of persons hospitalized, the length of a hospital stay is estimated to be 4 days [9, 10].

Additionally, for UK, we use the data provided by the *Office for National Statistics* (ONS)^2^. In particular, for the number of cases we use the daily estimates of infected population obtained by the Coronavirus (COVID-19) Infection Survey of the ONS. For the 7 days and the last month estimates, in order not to count multiple times the same cases, the sum of the daily percentages is divided by 10 days, an estimated average duration of the infection with Omicron [11]. Hospitalizations is the sum of the weekly admission rates with COVID-19 in England from Dec 19, 2022, to Jan 22, 2023 (5 weeks). Mortality is the rate of registered deaths involving COVID-19 in England from Dec 17, 2022, to Jan 20, 2023.

Finally, we use Cronbach’s Alpha coefficient to measure the reliability of the results obtained from the indirect surveys. Specifically, it quantifies the reliability of a value of an unobservable variable constructed from the observed variables. The closer this coefficient is to its maximum value of 1, the greater the reliability of the measure, but in general, it is considered that values greater than 0.7 are sufficient to guarantee reliability. There are several ways to calculate it, in this work the one based on the correlations of the observed variables is used.

## 3 Results

The comparison of our estimates with the official data in UK and Australia is presented in Table 1. The vaccination estimates are very close to the official values. The vaccination rates in Australia and UK are estimated as 76.50% (73.70% - 79.29%) and 78.86% (95% confidence interval: 77.00% - 80.72%), while the official (OWID) values are 84.95% and 79.71%, respectively. In the case of mortality and hospitalizations in the last month, the official values are within the confidence interval of our estimates in the case of Australia. Specifically, the mortality rate is 0.34% (0.00% - 0.22%) and the official is 0.006%, and the hospitalization rate is 1.02% (0.36% - 1.68%) and the official is 1.327%. Also, in the case of UK, the official values of ONS are within the confidence interval of our estimates of the number of cases, cases in the last 7 days, and cases in the last 24 hours.

For the rest of the variables, the differences are never abysmal in cases where there are major differences between the official values and our estimates (possibly due to underreporting in the official data). Cronbach’s alpha coefficient is 0.83 for Australia and 0.95 for the UK, which tells us that the reliability of the estimates is very good. The results of the estimates and Cronbach’s alpha coefficient allow concluding that we can use the indirect survey approach to make estimates when official data is not available or reliable, and use them considering a prudential bias when assessing them.

Table 2 shows the estimated results for China for all the questions of the survey. While 1.000 indirect survey responses were collected, the filters specified in Section 2.2 were used, reducing drastically the sample size to 469. Comparing our results with the OWID data for China, the vaccination rate is 91.9% while we estimate 91.03% (90.36%-91.7%), which is almost a perfect match. The estimate of the values for deaths is approximately 0.073% while we estimate 1.19% (0.94%-1.45%), a much higher value. However, OWID warns that “the number of confirmed deaths may not accurately represent the true number of deaths”. Therefore, our estimate could serve as a first approximation (that may be biased). Our estimate of number of cases in the last month is 78.57% (77.62%-79.54%), very far from 0.138% reported by OWID (which warns that “the number of confirmed cases is lower than the true number of infections”). Note that some areas of China may have a high incidence, as noted in the report published at [12]: “nearly 90% of Henan’s population had been infected by 6 January”.

**Table 2:** COVID-19 incidence metrics in % obtained from indirect survey data for China.

|  |  | Samples | Cases<br>(last month) | Vaccination<br>rate | Mortality<br>(last month) | Hosp<br>(last month) | Cases<br>(24 hours) | Cases<br>(7 days) |
| --- | --- | --- | --- | --- | --- | --- | --- | --- |
|  | **China** | 469 | **78.57**<br>**(77.62-79.54)** | **91.03**<br>**(90.36-91.70)** | **1.19**<br>**(0.94-1.45)** | **9.30**<br>**(8.61-9.97)** | **2.87**<br>**(2.48-3.26)** | **9.52**<br>**(8.83-10.21)** |
| Provinces | Jiangsu | 48 | 75.56<br>(72.42-78.69) | 87.92<br>(85.54-90.30) | 1.67<br>(0.73 - 2.60) | 7.64<br>(5.70-9.58) | 2.64<br>(1.47-3.81) | 9.44<br>(7.31-11.58) |
|  | Guangdong | 45 | 80.00<br>(76.98-83.02) | 86.07<br>(83.46-88.69) | 0.59<br>(0.01-1.17) | 5.33<br>(3.64-7.03) | 3.26<br>(1.92-4.60) | 6.96<br>(5.04-8.88) |
|  | Shandong | 27 | 74.81<br>(70.59 - 79.04) | 95.80<br>(93.85-97.76) | 1.48<br>(0.30-2.66) | 8.40<br>(5.69-11.10) | 2.22<br>(0.79-3.66) | 6.67<br>(4.24-9.10) |
| Cities | Shanghai | 9 | 68.89<br>(61.08-76.70) | 88.15<br>(82.70-93.60) | 2.22<br>(0.00-4.71) | 5.93<br>(1.94-9.91) | 0.74<br>(0.00-2.19) | 5.19<br>(1.44-8.93) |
|  | Guangzhou | 11 | 81.82<br>(75.93-87.70) | 86.67<br>(81.48-91.85) | 1.82<br>(0.00-3.86) | 9.70<br>(5.18-14.21) | 4.85<br>(1.57-8.13) | 7.27<br>(3.31-11.24) |
|  | Chengdu | 8 | 89.17<br>(83.61-94.73) | 88.33<br>(82.59-94.08) | 0.83<br>(0.00-2.46) | 8.33<br>(3.39-13.28) | 0.83<br>(0.79-2.45) | 8.33<br>(3.39-13.28) |
|  | Beijing | 8 | 74.17<br>(66.33-82.00) | 91.67<br>(86.72-96.61) | 0.83<br>(0.00-2.45) | 13.33<br>(7.25-19.42) | 5.00<br>(1.10-8.90) | 11.67<br>(5.92-17.41) |

We compute estimates for the provinces and cities with the largest number of samples (see Table 2). The rate of vaccination and cases in the last month is similar in all of them and similar to the values in China. The Guangdong province shows the lowest estimates of hospitalizations and deaths, while it has large case estimates among provinces. Among cities, Beijing shows low estimates of monthly cases, but large rates of recent cases and hospitalizations. Unfortunately, the sample size for cities is very small.

Finally, we would like to point out that, in general, the data are relatively small compared to the size of the country. Additionally, as can be seen in Table 3 in Supplementary Information, the sample is biased by age and education level. These biases are reduced with the use of indirect questions, but still more studies are needed.

**Table 3:** Characteristics of the survey respondents for Australia, the United Kingdom, and China.

| Characteristic | Australia | United Kingdom | China |
| --- | --- | --- | --- |
| 1. Number of participants | 100 | 200 | 1000 |
| 2. Gender, (%) |  |  |  |
| (a) Female | 56.00 | 58.00 | 46.90 |
| (b) Male | 44.00 | 42.00 | 53.10 |
| 3. Age groups, (%) |  |  |  |
| (a) 18-24 | 13.00 | 9.50 | 18.70 |
| (b) 25-34 | 27.00 | 26.00 | 44.30 |
| (c) 35-44 | 29.00 | 24.50 | 27.40 |
| (d) 45-54 | 17.00 | 22.50 | 8.40 |
| (e) >54 | 14.00 | 17.50 | 1.20 |
| 4. Education, (%) |  |  |  |
| (a) Middle school | 2.00 | 5.00 | 1.50 |
| (b) High school | 33.00 | 22.00 | 7.90 |
| (c) Technical college | 14.00 | 35.00 | 8.30 |
| (d) University | 43.00 | 25.00 | 63.30 |
| (e) Post-graduate | 7.00 | 11.50 | 18.90 |
| 5. Ethnicity, (%) |  |  |  |
| (a) Arab | 0.00 | 0.00 | 0.20 |
| (b) Asian | 8.00 | 7.50 | 94.60 |
| (c) Black | 0.00 | 2.50 | 0.20 |
| (d) Hispanic | 0.00 | 1.00 | 0.00 |
| (e) Latino | 0.00 | 0.00 | 0.20 |
| (f) White | 83.00 | 74.00 | 1.00 |
| (g) Multiracial | 3.00 | 1.00 | 0.20 |
| (h) Other | 6.00 | 14.00 | 1.60 |

## Data Availability

The data collected in the indirect surveys is publicly available at https://github.com/GCGImdea/coronasurveys/tree/master/papers/2023-COVID-19-China-January.

## Supplementary Information

### Questions of the Indirect Survey

#### Questions in English

Think of your 15 closest contacts in the last month. The rest of the questions below are with respect to this group of people. These contacts can be family, friends, or colleagues whose health status you know.

1. From the above 15 closest contacts in the last month, how many have had COVID-19 in the last month?
2. From the above 15 closest contacts in the last month, how many have been hospitalized for COVID-19 in the last month?
3. From the above 15 closest contacts in the last month, how many died from COVID-19 in the last month?
4. From the above 15 closest contacts in the last month, how many have COVID-19 today?
5. From the above 15 closest contacts in the last month, how many started with COVID-19 in the latest 7 days?
6. From the above 15 closest contacts in the last month, how many have (ever) been vaccinated for COVID-19?

#### Questions in Chinese

Contact the corresponding author to request access to this information.

## Acknowledgment

We want to thank Lin Wang for his help with the Chinese version of the survey.

## Footnotes

1 https://ourworldindata.org/

2 https://www.ons.gov.uk/

## References

[1] Astley Christina M, Tuli Gaurav, Mc Cord Kimberly A, et al. Global monitoring of the impact of the COVID-19 pandemic through online surveys sampled from the Facebook user base Proceedings of the National Academy of Sciences. 2021;118.

[2] Oliver Nuria, Barber Xavier, Roomp Kirsten, Roomp Kristof. Assessing the Impact of the COVID-19 Pandemic in Spain: Large-Scale, Online, Self-Reported Population Survey Journal of Medical Internet Research. 2020;22:e21319.

[3] Killworth Peter D, McCarty Christopher, Bernard H Russell, Shelley Gene Ann, Johnsen Eugene C. Estimation of seroprevalence, rape, and homelessness in the United States using a social network approach Evaluation review. 1998;22:289–308.

[4] Laga Ian, Bao Le, Niu Xiaoyue. Thirty years of the network scale-up method Journal of the American Statistical Association. 2021;116:1548–1559.

[5] Garcia-Agundez Augusto, Ojo Oluwasegun, Hernández-Roig Harold A, et al. Estimating the covid-19 prevalence in spain with indirect reporting via open surveys Frontiers in Public Health. 2021;9:658544.

[6] L. Cronbach. Coefficient alpha and the internal structure of tests Psychometrika. 1951;16:297–334.

[7] Hubert Mia, Vandervieren Ellen. An adjusted boxplot for skewed distributions Computational statistics & data analysis. 2008;52:5186–5201.

[8] Killworth Peter D, McCarty Christopher, Johnsen Eugene C, Bernard H Russell, Shelley Gene A. Investigating the variation of personal network size under unknown error conditions Sociological Methods & Research. 2006;35:84–112.

[9] Abdullah F, Myers J, Basu D, et al. Decreased severity of disease during the first global omicron variant covid-19 outbreak in a large hospital in tshwane, south africa International Journal of Infectious Diseases. 2022;116:38–42.

[10] Peralta-Santos André, Rodrigues Eduardo Freire, Moreno Joana, et al. Omicron (BA. 1) SARS-CoV-2 variant is associated with reduced risk of hospitalization and length of stay compared with Delta (B. 1.617. 2) MedRxiv. 2022:2022–01.

[11] Ries Julia. Omicron Infection Timeline: When Symptoms Start and How Long They Last Health. November 18, 2022.

[12] Lewis Dyani. China’s COVID wave has probably peaked, model suggests Nature. 2023;613:424–425.

